# An Explorative Assessment of ChatGPT as an Aid in Medical Education: Use it with Caution

**DOI:** 10.1101/2023.02.13.23285879

**Authors:** Zhiyong Han, Fortunato Battaglia, Abinav Udaiyar, Allen Fooks, Stanley R. Terlecky

## Abstract

**Objective:** To explore the use of ChatGPT by educators and students in a medical school setting.

**Method:** This study used the public version of ChatGPT launched by OpenAI on November 30, 2022 (https://openai.com/blog/chatgpt/). We employed prompts to ask ChatGPT to 1) generate a content outline for a session on the topics of cholesterol, lipoproteins, and hyperlipidemia for medical students; 2) produce a list of learning objectives for the session; and 3) write assessment questions with and without clinical vignettes related to the identified learning objectives. We assessed the responses by ChatGPT for accuracy and reliability to determine the potential of the chatbot as an aid to educators and as a “know-it-all” medical information provider for students.

**Results:** ChatGPT can function as an aid to educators, but it is not yet suitable as a reliable information resource for educators and medical students.

**Conclusion:** ChatGPT can be a useful tool to assist medical educators draft course and session content outlines and create assessment questions. At the same time, caution must be taken as ChatGPT is prone to providing incorrect information; expert oversight and caution are necessary to ensure the information generated is accurate and beneficial to students. Therefore, it is premature for medical students to use the current version of ChatGPT as a “know-it-all” information provider. In the future, medical educators should work with programming experts to explore and grow the full potential of AI in medical education.

## Introduction

The latest AI chatbot is ChatGPT (GPT stands for Generative Pre-trained Transformer) which is a large natural language model – that is, it interacts with users in a natural, conversational way and answers inquiries and follow-up questions from users. ChatGPT has been trained on text (approximately 300 billion words) available on the Internet and other open sources before the end of 2021, giving it the ability to respond to queries from users about virtually any topic (https://openai.com/blog/chatgpt/). Given that it can rapidly generate responses to user queries related to any information that it “read” during its training, ChatGPT offers myriad possibilities for experts as well as laypeople across our society.

The potential benefits of ChatGPT in teaching and learning is undeniable. The challenge for educators is how to creatively use ChatGPT as a tool. In one way, educators at universities have been testing the capability of ChatGPT in the absence of specialized training or reinforcement to answer questions in high-stake exams. It has been demonstrated that ChatGPT can pass the final exam of a typical Master of Business Administration core course with a “B” to “B-” grade on the exam.^1^ When tested using questions on the United States Medical Licensing Examination (USMLE), which consists of three exams: Step 1, Step 2CK, and Step 3, ChatGPT “performed at or near the passing threshold for all three exams.”^2^ Also ChatGPT performed, on average, at the level of a C+ student, achieving a low but passing grade on four law-school exams.^3^ In short, these examples suggest that ChatGPT has significant potential to assist teachers and learners – but much remains to be explored by the academic community.

Educators need to understand what ChatGPT is and is not. Like other large language models, ChatGPT was fed a gargantuan amount of text available on the Internet and other open sources before the end of 2021; ChatGPT was trained then to recognize the statistical probability of sequences of words that appear in this text (https://openai.com/blog/chatgpt/). The training resulted in ChatGPT being a statistical predictor that is capable of matching up strings of words that have statistical correlations to each other. Therefore, ChatGPT’s response to a prompt or a question from a user is entirely determined by what its algorithms deem to be the most statistically plausible response that resembles what it read in its training data. However, it should be emphasized that the text that was used to train ChatGPT contained inaccuracies and incorrect information - and that ChatGPT does not consider the truthfulness of its responses. Therefore, ChatGPT’s responses to prompts and inquiries from users are likely to contain inaccuracies and incorrect information. Another issue regarding ChatGPT is that it does not search for information from external data sources, and thus it produces responses based on information in its training text only. Therefore, the chatbot is likely to become confused and unable to provide quality responses to queries asking for up-to-date information. These issues raise serious concerns of how, and in what ways, educators and students can best use ChatGPT.

Nevertheless, it cannot be denied that ChatGPT is a very powerful tool and its wide use by medical educators and students, as well as teachers and learners in other fields, is inevitable. We ask here, in the context of medical education, should we embrace it or reject it? In this report, we describe our exploration of the potential of ChatGPT as an aid to medical educators as well as an information provider to medical students.

## Methods

This study used the public version of ChatGPT launched by OpenAI on November 30, 2022 (https://openai.com/blog/chatgpt/). We first evaluated diverse ways to converse with ChatGPT to obtain answers from ChatGPT to our questions in different formats regarding diverse biomedical concepts and issues. We also evaluated the ability of ChatGPT to write biomedical exam questions of different formats: short answers, essay type, and multiple-choice. We then used prompts to ask ChatGPT to 1) generate a content outline for a session on the topics of cholesterol, lipoproteins, and hyperlipidemia for medical students; 2) produce a list of learning objectives for the session; and 3) write assessment questions with and without clinical vignettes that are relevant to the learning objectives. We assessed accuracy and reliability of the information provided by ChatGPT to determine the potential of ChatGPT not only as an aid for educators but also as a “know-it-all” information provider for students.

When asking ChatGPT to write vignette-based multiple-choice questions, we were careful to suggest: using key words in the prompt to focus ChatGPT’s research on the most relevant information to the topic of the question; providing ChatGPT a context in the prompt for it to determine the level of detail; indicating the level of difficulty of the question (easy, difficult, challenging, very challenging) in the prompt.

## Results

### Using CHatGPT as an aid in the development of a session on cholesterol, lipoproteins, and hyperlipidemia

We asked ChatGPT what students should know regarding cholesterol, lipoproteins, and hyperlipidemia; and if it could outline the content of a session on the topic. For the content outline, the prompt we used was: Please outline the content of a session on cholesterol, lipoprotein, and hyperlipidemia for medical students. Within seconds, ChatGPT produced the following content outline.

1. Introduction to cholesterol and lipoproteins
  - Definition of cholesterol and lipoproteins
  - The different types of lipoproteins (LDL, HDL, VLDL, IDL) and their roles in the body
  - The normal range for cholesterol and lipoprotein levels
2. Causes and risk factors for high cholesterol
  - Genetic factors
  - Diet and lifestyle factors (such as high intake of saturated and trans fats, lack of physical activity, and smoking)
  - Medical conditions (such as diabetes and hypothyroidism)
3. Hyperlipidemia
  - Definition of hyperlipidemia and its subtypes (primary and secondary)
  - The link between hyperlipidemia and cardiovascular disease
  - The importance of early diagnosis and treatment
4. Measuring cholesterol and lipoprotein levels
  - Different methods of measuring cholesterol and lipoprotein levels (such as fasting lipid profile and non-fasting lipid profile)
  - How to interpret test results and what they mean
5. Treating high cholesterol
  - Lifestyle changes (such as a healthy diet, regular exercise, and weight management)
  - Medications (such as statins, fibrates, and niacin)
  - Surgery (such as LDL apheresis and bariatric surgery)
  - The importance of monitoring and follow-up
6. Guidelines for screening and treating high cholesterol
  - National guidelines for screening and treating high cholesterol (such as the Adult Treatment
  - Panel III and the American College of Cardiology/American Heart Association guidelines)
  - Special considerations for different populations (such as children, women, and older adults)
7. Case studies and discussions
  - Review of case studies of patients with high cholesterol and hyperlipidemia
  - Discussions on how to approach diagnosis and treatment in these cases.
8. Conclusion
  - The importance of understanding cholesterol, lipoproteins, and hyperlipidemia in the practice of medicine
  - The importance of early diagnosis, treatment, and follow-up for reducing the risk of cardiovascular disease

The outline is of decent quality, but it is puzzling that ChatGPT listed “surgery (such as LDL apheresis and bariatric surgery)” as a treatment option for patients with high cholesterol. The use of “surgery” is problematic because bariatric surgery is not used for patients with high cholesterol and LDL apheresis is a nonsurgical treatment. ChatGPT is correct in that LDL apheresis is used for patients with high LDL cholesterol levels, especially those with familial hypercholesterolemia.^4^ Therefore, here we would change “Surgery (such as LDL apheresis and bariatric surgery)” to Lipoprotein apheresis.

We then asked ChatGPT to create a list of learning objectives for this session; the prompt used was, “Based on the outlined content, please write out a list of session learning objectives.” Again, within seconds, ChatGPT produced the following learning objectives.

1. Explain the definition, composition and roles of cholesterol and lipoproteins in the body.
2. Describe the normal range for cholesterol and lipoprotein levels and explain the significance of each.
3. Identify the causes and risk factors for high cholesterol and hyperlipidemia.
4. Explain the link between hyperlipidemia and cardiovascular disease and the importance of early diagnosis and treatment.
5. Describe the different methods of measuring cholesterol and lipoprotein levels and how to interpret test results.
6. Outline the various treatment options available for managing high cholesterol, including lifestyle changes, medications, and surgery.
7. Explain the importance of monitoring and follow-up in managing high cholesterol.
8. Describe national guidelines for screening and treating high cholesterol, and special considerations for different populations.
9. Analyze case studies of patients with high cholesterol and hyperlipidemia to apply knowledge and concepts learned in the session.
10. Develop an understanding of the importance of understanding cholesterol, lipoproteins, and hyperlipidemia in the practice of medicine.

These learning objectives are adequate but do require some revision. Interestingly, ChatGPT included “surgery” in learning objective 6, which is incorrect as we have already discussed. Moreover, ChatGPT left out an important learning objective – that is, describe the metabolism of cholesterol and lipoproteins and how each process is regulated by specific enzymes. This information is critical for students to understand the distinct types of completely hyperlipidemia, the mechanism of action of various anti-hyperlipidemic drugs, the development of such agents, and the diagnosis, treatment and management of patients with hyperlipidemia. In this example, ChatGPT did not always “think” like a trained medical educator.

Next, we instructed the chatbot - “Please write multiple-choice questions with a vignette and containing lab values”. Below is an example of what was produced.

A 35-year-old woman presents with a history of obesity and type 2 diabetes. She is found to have a triglyceride level of 800 mg/dL and an HDL level of 25 mg/dL. What is the most likely cause of her hyperlipidemia?

A. Familial hypertriglyceridemia
B. Familial combined hyperlipidemia
C. Metabolic syndrome
D. Secondary hyperlipidemia

The correct answer is C. Metabolic syndrome. This is a reasonable basic question to assess students’ knowledge of the diagnostic criteria for metabolic syndrome, which include obesity, type 2 diabetes, elevated triglyceride levels and low HDL levels.^5^ Next, we asked ChatGPT to improve this question by adding more lab values; the revised question is below.

A 35-year-old woman presents with a history of obesity and type 2 diabetes. She is found to have a triglyceride level of 800 mg/dL, an HDL level of 25 mg/dL, a fasting glucose level of 126 mg/dL, and a blood pressure of 140/90 mmHg. What is the most likely cause of her hyperlipidemia?

A. Familial hypertriglyceridemia
B. Familial combined hyperlipidemia
C. Metabolic syndrome
D. Secondary hyperlipidemia

The inclusion of fasting glucose and blood pressure helps to provide a more complete picture of the patient’s metabolic status and can help to increase the accuracy of the diagnosis. That said, we would have included more in the vignette, including information about lifestyle, body mass index, and waste circumference – parameters commonly considered in conjunction with metabolic syndrome.^5^

To further test ChatGPT, we used the prompt “Can you change this question into one for which familial combined hyperlipidemia is the best answer?” Below is the updated question and ChatGPT’s explanation.

A 35-year-old woman with a positive family history of hyperlipidemia presents with elevated levels of total cholesterol, LDL cholesterol, and triglycerides. Her lipid panel shows a total cholesterol level of 270 mg/dL, an LDL cholesterol level of 180 mg/dL, and a triglyceride level of 500 mg/dL. Her HDL level is 35 mg/dL. What is the most likely cause of her hyperlipidemia?

A. Familial hypertriglyceridemia
B. Familial combined hyperlipidemia
C. Metabolic syndrome
D. Secondary hyperlipidemia

“In this revised question, the presence of elevated levels of total cholesterol, LDL cholesterol, and triglycerides, in addition to a positive family history of hyperlipidemia, supports the diagnosis of Familial combined hyperlipidemia, which is characterized by elevated levels of multiple lipid fractions.”

The values of LDL cholesterol, HDL cholesterol, and triglycerides in the vignette fits with familial combined hyperlipidemia, and the explanation is reasonable. However, we would have included an elevated serum ApoB value in the scenario because it is highly relevant to the diagnosis and prognosis of patients with this disorder.^6^ To further improve the question, we would replace “a positive family history of hyperlipidemia” in the vignette with the following: “a family history of premature cardiovascular disease” - to indicate an important *clinical* feature of the disease.^7,8^

### Can ChatGPT be a useful “know-it-all” information provider for medical students?

One of the major changes in medical education in recent history is the transformation of the traditional curriculum to an integrated curriculum (also known as organ- or system-based curriculum).^12^ The traditional approach teaches students each discipline of the basic sciences and clinical medicine in distinct silos, with the result that students struggle to make sense of the connection between basic science content (e.g., anatomy, cell biology, biochemistry, immunology, microbiology, nutrition, pharmacology, and physiology) and the disease state. In contrast, an integrated curriculum aims to connect basic sciences and clinical medicine in ways that permit students to see the relevance of basic science to patients and medicine. Such integrate curricula allows students to develop a clear understanding of each organ/system in healthy and disease states - with the underlying molecular and cellular underpinnings carefully presented and considered.^9^ This knowledge is critical for physicians to be able to lead the development of drugs, therapeutics, and diagnostic tests for disease. Additionally, an integrated curriculum aims to better prepare students to acquire critical analytic and reasoning skills to provide the ability to solve complex medical problems and to create life-long, self-dependent learning capabilities.^9^ One of the ultimate goals of teaching students in an integrated curriculum is to produce clinicians whose thought process regarding the diagnosis and treatment of disease is no longer demarcated by a “boundary” between basic sciences and clinical medicine.^9^

One of the issues related to both faculty teaching and student learning in an integrated curriculum concerns its compressed nature. Often, the length of the curriculum is 14-16 months instead of the more traditional two years. Compressed preclinical curricula discourage didactic teaching by faculty because such traditional approaches (e.g., lectures) promote passive learning by students. Modern pedagogical approaches encourage active learning by students. Active learning requires students to resolutely engage with the course material all the time. Moreover, it requires a significant amount of self-directed study of the course materials often in the absence of faculty guidance.^10–12^ The advantage of active learning is that it promotes a deeper understanding of the material and requires students to master and apply both basic sciences and clinical medicine to explain the pathophysiology of specific disease. Furthermore, it necessitates students develop higher cognitive skills and the ability to integrate knowledge and transfer same in new contexts in which content application and synthesis are required.

Given the requirement for active learning by students while they encounter enormous amounts of scientific material, it is inevitable that many students experience stress,^11^ and active learning curriculum can negatively impact students’ attitudes towards active learning because they have not acquired sufficient knowledge and skill to be active learners.^12^

Students may consider ChatGPT a polymath and an expert in every discipline; therefore ChatGPT may very well be an ideal “know-it-all” information provider to students anytime and anywhere because asking ChatGPT questions is akin to asking professors questions without the concern of being wrong, appearing unprepared, or feeling embarrassed. However, as emphasized by ChatGPT - it is a text-based AI model and does not have the capability to show, study, and create images. Therefore, ChatGPT may not be an ideal for visual learners, and it certainly is not a tool for students to learn anatomy, histology, and imaging techniques. Nevertheless, what we learned subsequently from ChatGPT gave us serious pause about the chatbot and its ability to teach students. Specifically, ChatGPT provided factually incorrect information about lipoproteins. Consider: ChatGPT stated, “LDL [low-density lipoprotein] is synthesized in the liver ….” This is simply wrong because LDL is a catabolic product of VLDL (very low-density lipoprotein) in the capillaries of extrahepatic tissues and VLDL is synthesized in hepatocytes.

ChatGPT stated, “The RCT [reverse cholesterol transport] process begins in peripheral tissues such as the arterial wall, where excess cholesterol is taken up by a cell surface receptor called scavenger receptor class B type I (SR-BI). The cholesterol is then transported to the liver by an enzyme called ATP-binding cassette transporter A1 (ABCA1).” The fact is that ABCA1 transports intracellular cholesterol to the outside the cells in extrahepatic tissues, and the cholesterol is then taken up by HDL in a process involving the action of an enzyme called LCAT (lecithin cholesterol acyltransferase). Finally, HDL interact with SR-BI in the membrane of hepatocytes to transfer its cholesteryl ester content inside.^13^

ChatGPT stated, “On the other hand, individuals with genetic mutations or variations that lead to reduced CETP activity have been found to have higher levels of HDL-cholesterol and lower levels of LDL-cholesterol and have a lower risk of cardiovascular diseases.” The fact is that the risk of cardiovascular disease associated with mutations in CETP is more complicated.^14^

After seeing the statement by ChatGPT regarding the cardiovascular risk associated with CETP mutations, we asked ChatGPT a question: “Some mutations in CETP seem to increase the risk of cardiovascular diseases. So, what is going on?” ChatGPT provided a revised statement:

“Some genetic mutations in the CETP gene have been found to increase the risk of cardiovascular diseases. These mutations lead to a reduced activity of CETP, which results in increased levels of HDL-cholesterol and decreased levels of LDL-cholesterol. However, this does not mean that these mutations are always beneficial. One of the best-known CETP mutations is the I405V mutation, which is associated with increased CETP activity and decreased levels of HDL-cholesterol. Individuals with this mutation have been found to have a higher risk of cardiovascular diseases, such as coronary artery disease, than individuals without the mutation. Another mutation is the T-Insertion, which is associated with decreased CETP activity and increased levels of HDL-cholesterol. Individuals with this mutation have been found to have a lower risk of cardiovascular diseases, such as coronary artery disease, than individuals without the mutation. It’s important to note that these findings are not conclusive and that other factors such as lifestyle, diet and overall health status may also play a role in the development of cardiovascular diseases. In summary, some genetic mutations in the CETP gene have been found to be associated with increased or decreased risk of cardiovascular diseases. These mutations lead to changes in the activity of CETP and the levels of lipoprotein, such as HDL-cholesterol and LDL-cholesterol. However, it’s important to note that these findings are not conclusive and that other factors such as lifestyle, diet and overall health status may also play a role in the development of cardiovascular diseases.”

We asked ChatGPT for references regarding the I405V and the T-insertion mutations; it provided 5 references with titles and lead author names. However, we could not verify these citations. Nevertheless, the truth is that the relationship between the mutations in CETP and cardiovascular risk remains unclear with results often depending on study methodologies and/or other factors.^14^

We have found another issue concerning the use of ChatGPT as a “know-it-all” information provider; specifically, its explanations of how things work lacks depth and sometimes contains incorrect information. For example, we gave ChatGPT the prompt, “Please describe what PCSK9 is and its mechanism of action”; ChatGPT produced the following:

“PCSK9 (Proprotein Convertase Subtilisin/Kexin type 9) is a protein that plays a role in regulating the levels of low-density lipoprotein (LDL) cholesterol in the blood. Specifically, PCSK9 binds to LDL receptors on liver cells, which normally help to remove LDL cholesterol from the bloodstream. When PCSK9 binds to these receptors, it causes them to be degraded rather than recycled, thereby reducing the number of receptors available to remove LDL cholesterol. This results in a rise in LDL cholesterol levels in the blood, which is a risk factor for cardiovascular disease. Inhibitors of PCSK9, called PCSK9 inhibitors, work by blocking the activity of PCSK9 and thereby increasing the number of LDL receptors available to remove LDL cholesterol from the blood, resulting in a decrease of LDL cholesterol levels.”

At first glance, what ChatGPT provided is not wrong. But it does not provide the mechanism of action of PCSK9. Therefore, we asked ChatGPT with a follow-up prompt, “Please describe the mechanism of action of PCSK9.” This time, ChatGPT provided more information - see below.

PCSK9 is a protein that plays a role in regulating the levels of low-density lipoprotein (LDL) cholesterol in the blood. The mechanism of action of PCSK9 is as follows:

1. PCSK9 is produced by the liver and circulates in the bloodstream.
2. PCSK9 binds to LDL receptors on liver cells. LDL receptors are responsible for binding to LDL cholesterol particles in the blood, and then internalizing and degrading them.
3. When PCSK9 binds to LDL receptors, it causes the receptors to be degraded instead of recycled. This reduces the number of receptors available to remove LDL cholesterol from the bloodstream.
4. As a result, there are fewer LDL receptors available to remove LDL cholesterol from the blood, leading to an increase in LDL cholesterol levels. This is a risk factor for cardiovascular disease.

However, the information is still insufficient. Therefore, we asked another guiding question: “Does the lysosome play a role in this process?” ChatGPT responded:

“Lysosomes do play a role in the process of PCSK9 regulating LDL cholesterol levels. When an LDL receptor binds to an LDL cholesterol particle, the LDL receptor-LDL cholesterol complex is internalized and transported to a lysosome. Once inside the lysosome, the LDL cholesterol particle is degraded by lysosomal enzymes. However, in the presence of PCSK9, the LDL receptor-PCSK9 complex is also internalized, but it is not delivered to the lysosome for degradation. Instead, it is targeted for degradation in the endosome. This process leads to the downregulation of LDL receptors and reduction in the number of LDL receptors that are available for binding and internalizing LDL cholesterol particles. In summary, lysosomes play a role in degradation of LDL cholesterol particles after they have been internalized by LDL receptors. But the process of PCSK9 binding to LDL receptors and downregulating the number of available receptors does not directly involve lysosomes.”

This response is full of incorrect information, and it would have fooled students who just started learning this topic and even educators who are not well versed in lipoprotein metabolism. First, it forgot to mention that the LDL receptor in the internalized LDL-LDL receptor complex is normally recycled back to the cell surface.^15^ Second, the internalized LDL-LDL receptor complex is routed to endosomes where segregation of LDL and LDL receptors occurs, and the LDL receptor is recycled back to the cell surface whereas LDL is routed to the lysosome to be degraded.^15^ Third, PCSK9-facilitated degradation of LDL receptor does not occur in the endosome but rather in the lysosome.^16,17^ To test ChatGPT further, we asked it this question: “Are LDL receptors recycled back to cell surface in the absence of PCSK9 action?”

“Yes, LDL receptors are recycled back to the cell surface in the absence of PCSK9 action. The LDL receptors are internalized when they bind to LDL cholesterol particles, and in the absence of PCSK9, the LDL receptor-LDL cholesterol complex is delivered to a lysosome for degradation. Once inside the lysosome, the LDL cholesterol particle is degraded by lysosomal enzymes and the LDL receptor is recycled back to the cell surface. The recycled LDL receptors can then bind to and internalize more LDL cholesterol particles, leading to a continuous removal of LDL cholesterol from the bloodstream. In the presence of PCSK9, the LDL receptor-PCSK9 complex is also internalized, but it is not delivered to the lysosome for degradation. Instead, it is targeted for degradation in the endosome, this process leads to downregulation of LDL receptors, reduction in the number of LDL receptors that are available for binding and internalizing LDL cholesterol particles, which results in an increase in LDL cholesterol levels.”

This time, ChatGPT agreed that the LDL receptor can be recycled back to cell surface, but it got it wrong about where rerouting of the LDL receptor to the cell surface starts. Also, it makes the same mistake identified above by stating that PCSK9-facilitated degradation of LDL receptor occurs in the endosome.

## Discussion

In this explorative study, we have learned that ChatGPT sometimes offers factually inaccurate answers to basic science questions and can provide misleading medical information. As but one example, ChatGPT tried, and failed, to correctly link CETP mutations and cardiovascular disease risk and the relationship among LDL receptors, endosome, and lysosome in the context of LDL receptor recycling in the absence and presence of PCSK9. It appears that ChatGPT can simply produce statements that its algorithms deem to be the most statistically plausible-sounding responses to questions from users without regard to accuracy. Identification and correction of such inaccuracies require knowledgeable and astute human medical experts.

It is well known that writing high-quality multiple-choice questions is a difficult and time-consuming task for educators.^18,19^ Although some of the questions generated by ChatGPT may not be useful as they are, the fact that chatbox can rapidly create vignette-based multiple-choice questions concerning a topic means educators are provided “raw materials” to work with. That is, educators may edit/revise the questions to ensure that they do not present inaccurate information and that they truly reflect what was presented in learning sessions and expected to be understood. Therefore, using ChatGPT would decrease the time and effort required for educators to write examination questions. Additionally, it suggests that educators can use ChatGPT to generate large question banks for students to use for self-assessment of their learning. One could envision encouraging medical school faculty members to collaborate with each other to use ChatGPT and develop customized question banks that are specific for their courses and session objectives. However, the key to development of high-quality question banks is that the team carefully review, edit, and revise ChatGPT-generated questions to assure that the final questions are accurate and truly appropriate for the student learning assessments.

Clearly, students need to be informed that although ChatGPT in its current form seems to provide useful information relevant to the material they are learning, they need to understand know that they may receive superficial information and that the chatbot can provide inaccurate results. The problem, of course, is that students may be unable to discern such bogus information due to their lack of a sophisticated and deep understanding of the content material. To acquire correct and in-depth information from ChatGPT requires the active learning capability of users because only those who can identify their knowledge gaps and understand what they do not know can use ChatGPT efficiently. Unfortunately, many medical students in a preclinical curriculum are still developing such active learning capabilities. Therefore, medical students should not be too eager to embrace and use ChatGPT as their “know-it-all” information provider at the present time.

In conclusion, ChatGPT can be a useful tool to assist medical educators to draft course and session content and create materials for the development of valuable question banks. At the same time, caution must be taken as ChatGPT is prone to providing incorrect information regarding some topics and is not meant to be a reliable source of information for both educators and students. For those students who use ChatGPT as a source of information, expert oversight and caution are necessary to ensure the information generated is accurate and beneficial to students.

## Data Availability

All data produced in the present work are contained in the manuscript

## Contributions

ZH, FB, and SRT are responsible for conceiving the study and writing the manuscript; ZH and BF were involved in exploring ChatGPT’s potential; AU and AF also explored ChatGPT’s potential and were involved in manuscript writing.

